# An Ecological Study on the Associations of County-Based Credit Ratings with Age-Adjusted Mortality Rates in Michigan

**DOI:** 10.1101/2023.04.08.23288293

**Authors:** John P Rogers, Tyler J Dempsey

**Affiliations:** Michigan State College of Osteopathic Medicine

## Abstract

The objective of this study was to examine the association and implications between 21 Michigan county-specific credit ratings and age-adjusted mortality rates averaged across six quarters of county-specific data. Pearson’s regression analysis was performed and indicated that county credit ratings were significantly associated with average age-adjusted county mortality rates (r = 0.7715, p = 0.0000421), with lower county credit ratings corresponding to higher average age-adjusted county mortality rates. The results indicate that county credit ratings are associated with negative public health outcomes specific to that county, which the investigators propose mechanistically relates difficulty of local governments ascertaining low-cost funding for community health, infrastructure, and economic development projects.

## Introduction

Socioeconomic disparity has been increasingly focused on in recent years, at least in part due to the disparity of health outcomes during the COVID-19 pandemic. Economically, monetary policy continues to tighten; with rising interest rates disproportionately hurting lower rated borrowers. In light of this, a holistic approach to public health and economics research is increasingly needed. Interdisciplinary research is key to unlocking better policy guidance in order to narrow inequalities at the local level.

County mortality rates are an important metric to judge the general health of local communities. While county mortality rates are, of course, multivariate, numerous studies have correlated the socioeconomic health and demographics of counties to increased county mortality rates^1-3^. In addition, negative health outcomes have been associated with financially burdened individuals within a community^4-7^.

One study by <u>Adepu et. al</u>. in Georgia found a statistically significant negative correlation between cardiovascular disease mortality and income on a county level^8^. Another study by <u>Gong et. al</u>. used a wellbeing index composed of 10 socioeconomic factors, with a higher wellbeing index meaning higher socioeconomic disparity, and compared this to county mortality rates. This study found a significant positive association between the two variables^**9**^.

Another angle to look at socioeconomic health is credit ratings within the local community of study. Lower personal credit ratings have been shown to be associated with both lower self-reported health and higher rates of cardiovascular disease^10,11^. While individual’s credit ratings have been assessed in the literature, we identified a gap pertaining to broader government credit ratings and their associations with, and effects on, health outcomes within communities.

Therefore, we decided to look into county government measures of socioeconomic health and their relation to county public health outcomes. Specifically, we thought county government credit ratings would be a good metric to assess the socioeconomic health of a county, and county mortality rates would be a good public health outcome to look at. Since to date no one has attempted to study this, *we hypothesized that lower Michigan county credit ratings will predict higher age-adjusted county mortality rates*.

### Background on County Credit Ratings

County government credit ratings are, in general, a function of the county’s bonds in circulation. Bonds can be issued by County, State and National Governments to fund government expenses. Local governments, such as county governments, issue bonds to cover large expenses, and these bonds are known as “Municipal Bonds’’. Such municipal bonds may be issued for large expenses such as a new public library, sewage system modernizations, jail construction, etc.^12^.

Credit rating agencies are sometimes recruited to assign ratings involving letters and numbers to these newly-issued bonds. The assigned rating corresponds to the health of the bond issuer. For instance, an Aa1 rated bond would mean the bond issuer is extremely unlikely to default on the loan, whereas a Ba1 rated bond would mean there is substantial risk involved for the bond purchaser (See Figures 1 & 2).

**Figure 1.**
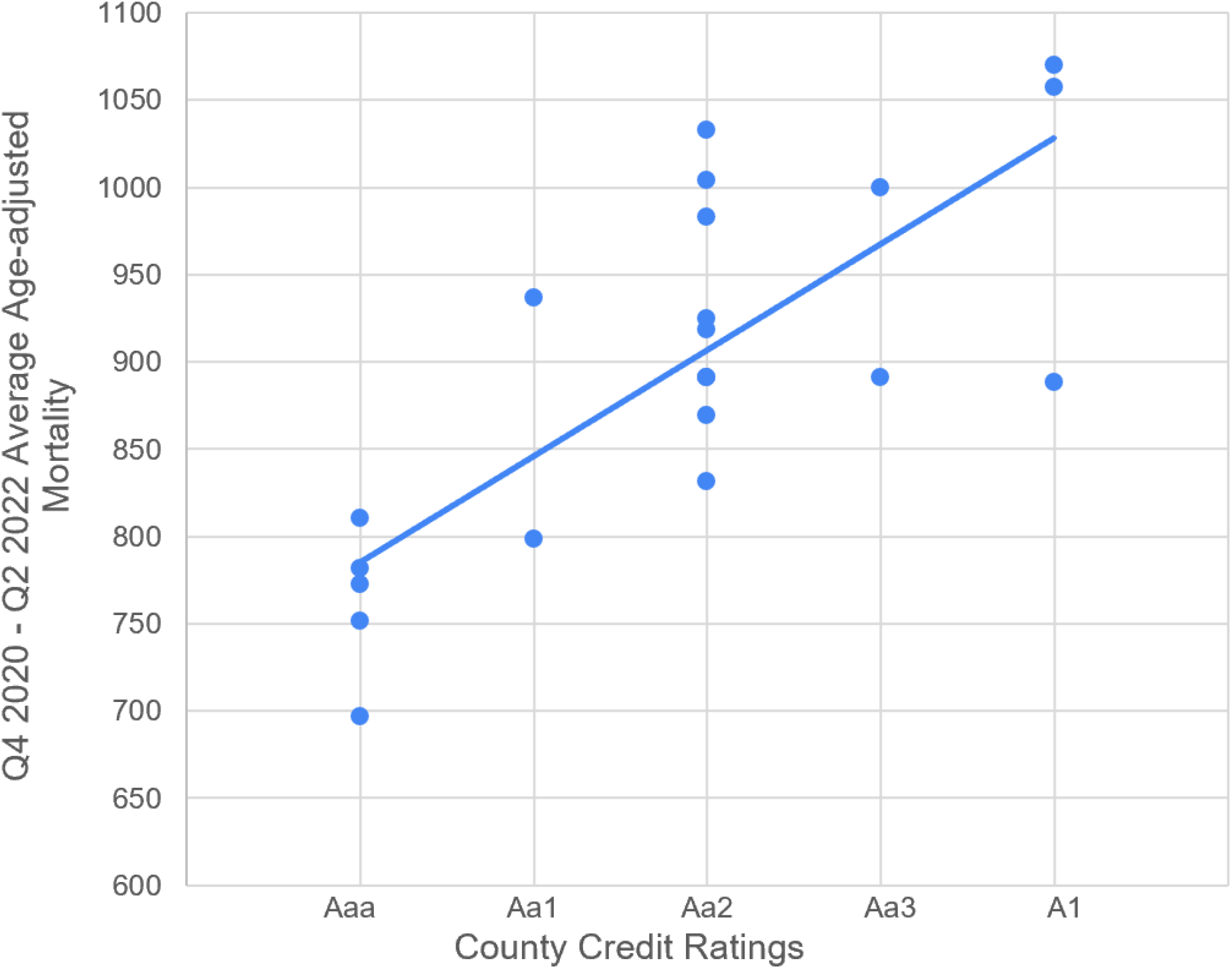
Graph of relationship between County Credit Ratings and Average Age- adjusted County Mortality Rates.

**Figure 2.**
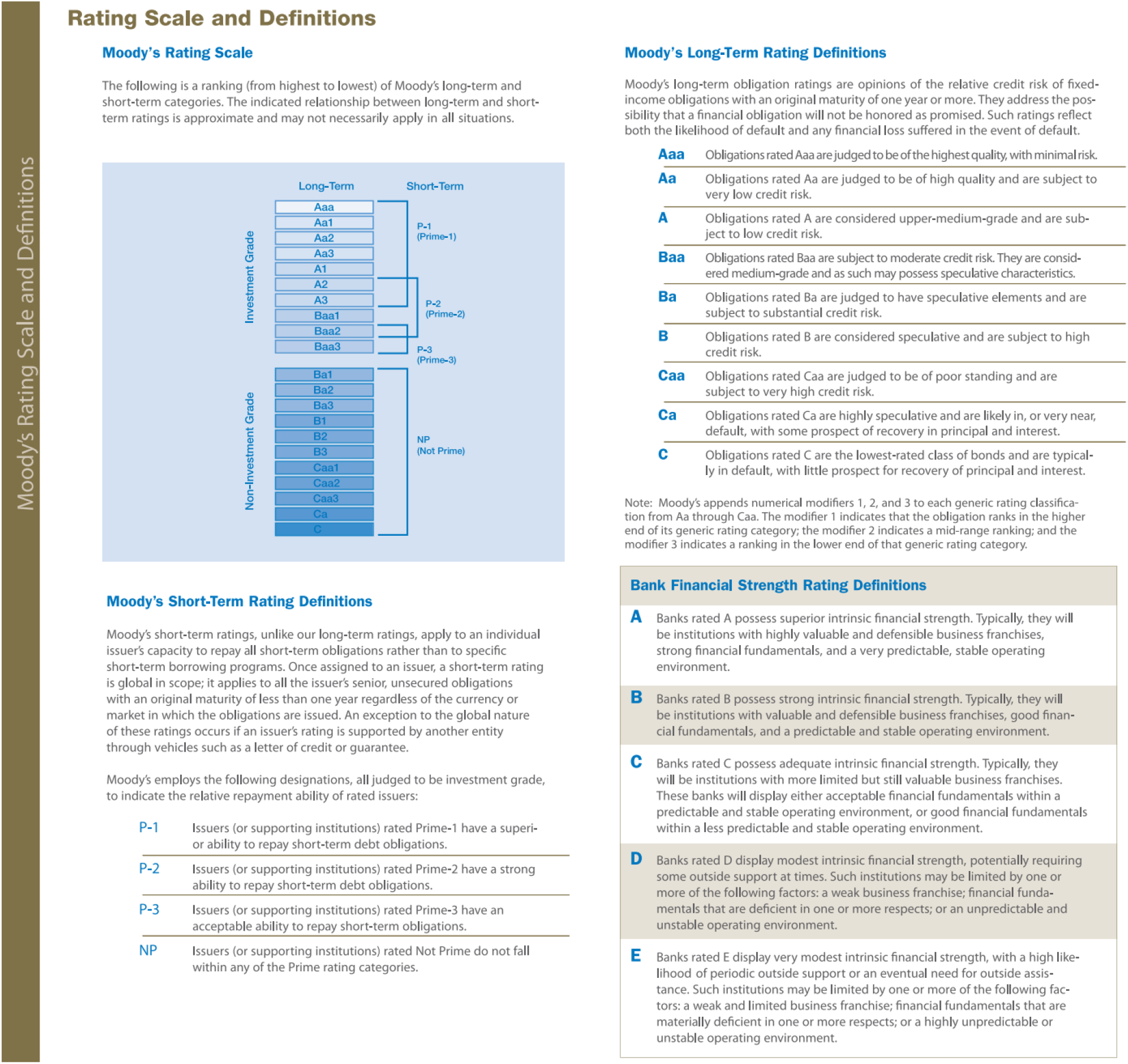
Moody’s Rating Scale with Definitions. Source - https://www.moodys.com/sites/products/productattachments/ap075378_1_1408_ki.pdf

As stated above, these bonds are typically rated by a bond rating agency, such as Moodys, Fitchs, or Standard and Poors Global. Local governments such as counties are typically assigned lower credit ratings based on a slew of ratings criteria, many of which give a window into the socioeconomic health of a specific county. Socioeconomic factors included in the ratings criteria include: lower tax revenues, lower median family income, lower manufacturing index scores (MFI), higher indebtedness, and lower cash balances^13,14,^ Figure3. Moody’s, the rating issuer in this study, provides a “scorecard” methodology to show how they guide local government ratings. This scorecard breakdown is summed up in Figure 3.

**Figure 3.**
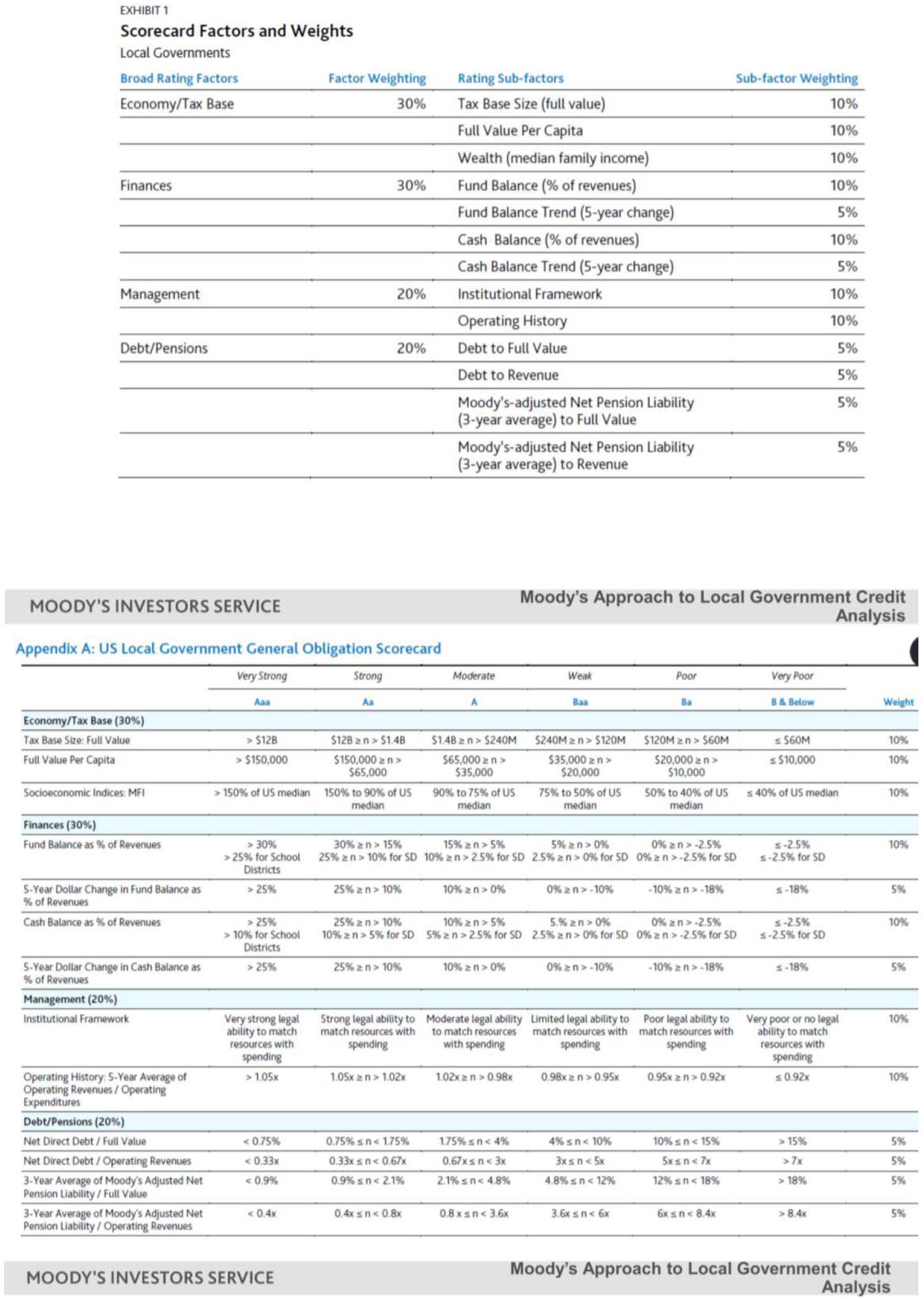
Moody’s Scorecard for Local Government Bond Ratings. Source - https://www.treasurer.ca.gov/cdiac/seminars/2019/20190212/day2/6.pdf

Municipal ratings can therefore be used as a general measure of the economic health of the local government who issues them. Lower ratings therefore correspond to higher risk of the local government defaulting on the loan, and ratings directly influence the interest rate on the bond, with lower ratings meaning higher interest rates.

## Methods

### Study Design

In order to answer the question of whether lower Michigan county credit ratings will predict higher age-adjusted county mortality rates, we performed a cross-sectional analysis of 21 Michigan counties over 6 quarterly intervals, with those being Q4 2020 to Q1 2022.

### County Credit Ratings and Selection

We collected publicly available ratings data from Moody’s (a bond rating agency) on all current Michigan county-specific credit ratings^**15**^. Note that only counties with active Moody’s ratings were pulled for analysis in order to ensure information was current. Of the 83 counties in Michigan, only 21 counties had active ratings and were subsequently included in the analysis, with bond ratings ranging from the Aaa (highest) to A1 (lowest in Michigan) (See Figure 2). Note that we excluded ratings from Fitch, as few active county ratings were found, and no active county credit ratings were found from Standard & Poors Global.

### Mortality Rates

Quarterly age-adjusted mortality rates by 12-month ending were obtained from the Michigan Department of Health and Human Services (MDHHS)^16^. Using age-adjusted data allowed us to control for the large variations in county population. Crawford county was the smallest, with a population around 13,000 persons, and Wayne county was the largest, with around 1.8 million persons.

Six quarters of all-cause county specific mortality rates by 12-month ending were included in the data, covering a period from Q4 of 2020 to Q1 of 2022. This quarterly data was then averaged across the six quarters in order to look at the overall trend over the study period. This design was chosen due to the county credit ratings holding steady over the study’s time period (Q4 2020 - Q1 2022), allowing us to better align the relationship between a county’s credit rating and its age-adjusted mortality rate.

### Statistical Methods

Linear regression analysis was used to determine the correlation coefficient. Analysis was performed in Microsoft Excel 2016 with a 95% confidence interval (alpha of 0.05) to determine statistical significance.

## Results

Regression analysis was performed to examine the relationship between Michigan county-specific credit ratings and average age-adjusted Michigan county mortality rates. The findings indicated that county credit ratings were significantly positively associated with average age-adjusted county mortality rates (r = 0.7715, p = 0.0000421), with lower county credit ratings corresponding to higher average age-adjusted county mortality rates. This can be seen in Figure 1 below.

The age-adjusted county mortality rates ranged from 810.2 deaths per 100,000 persons (Kent county) to 1057.12 deaths per 100,000 persons (Wayne county). Analysis showed that for every 1 decrease in bond-ratings (i.e. from Aaa to Aa1), there was an associated 60.75±11.49 increase in age-adjusted county mortality rates per 100,000 persons.

## Discussion

The findings of this study show that there is a significantly positive relationship between county credit ratings and age-adjusted mortality rates in Michigan. Counties having lower credit ratings tended to have higher age-adjusted mortality rates, on average, compared to counties with higher credit ratings. Using age-adjusted mortality rates controlled for population size as a potential confounding variable.

The findings suggest that credit ratings of a specific county could be a predictor of public health outcomes in Michigan. The potential reasons for this are complex and multifactorial. Our proposed mechanism described below can be found in the form of a flowchart provided in Figure 4.

**Figure 4.**
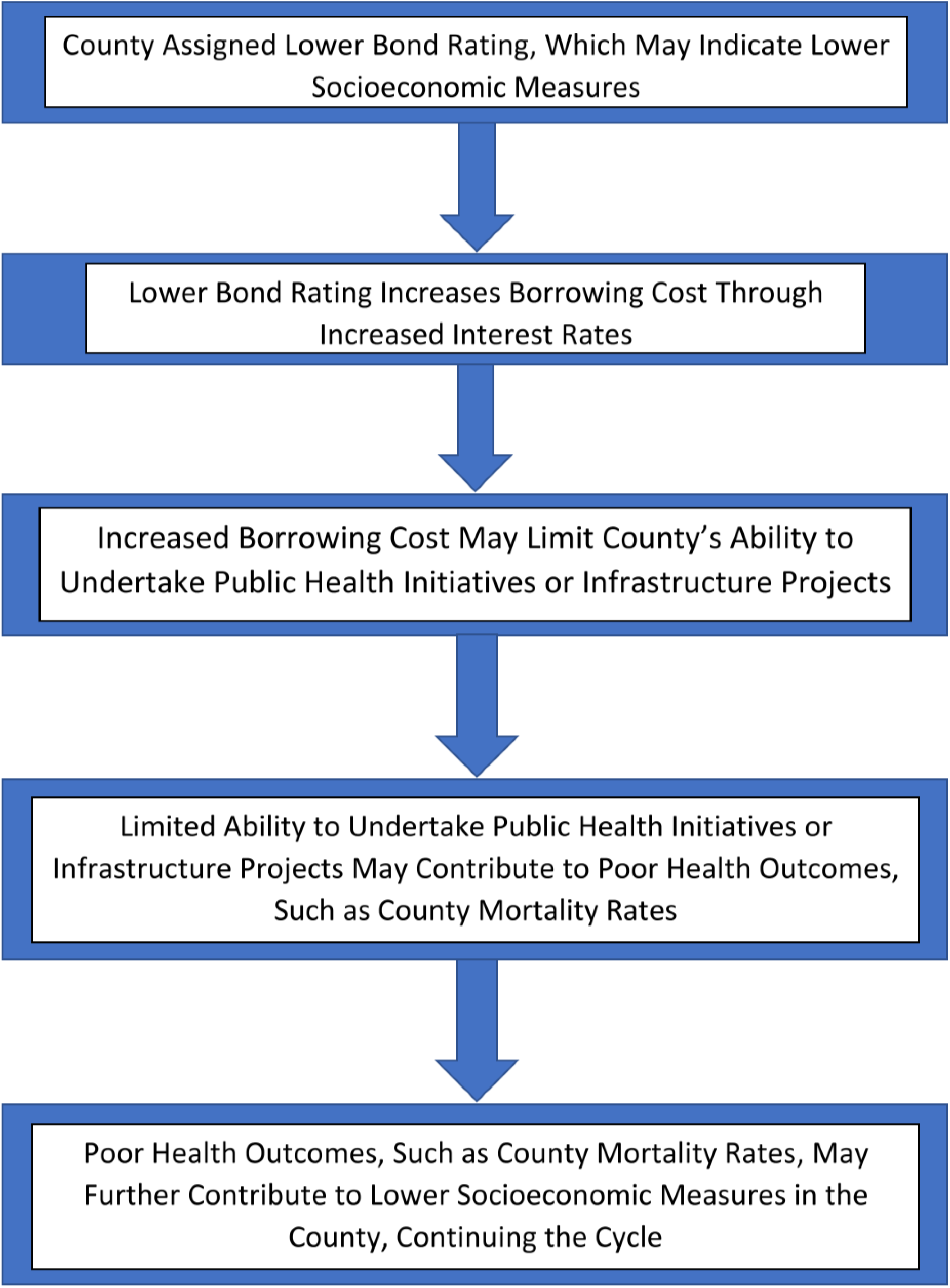
Proposed mechanism of how lower bond ratings may affect mortality rates.

As stated in the introduction, lower credit ratings for county governments may be reflective of generally lower socioeconomic measures in a county. With lower credit ratings, higher bond interest rates are seen. Higher interest rates means subsequently higher borrowing costs.

With higher borrowing costs, the ability for a county to undertake public health initiatives or infrastructure projects may be mitigated, or larger deficits must be taken on^17,18^. Some examples of public health-related Michigan county projects funded by municipal bonds include: Hospital revenue and refunding, economic development loans to companies, sewer disposal system refunding, and refuse disposal system refunding^19^.

Fewer public health initiatives and/or infrastructure projects due to high borrowing costs may contribute to poor health outcomes. This could be due to a variety of factors, but using the examples from above would include: reduced access to resources (lack of funding for a hospital), reduced sanitation (in the case of sewage and water systems bonds), or lack of funding for economic development contributing to lower socioeconomic status of the community.

Taken together, this lack of public health initiatives and infrastructure projects could contribute to poor health outcomes in the county. As discussed in the introduction, reduced socioeconomic health and mortality rates have been shown to have strong associations.

According to the factors discussed above, as well as the ratings criteria discussed in the introduction, a more robust picture of county credit ratings as a function of socioeconomic disparity emerges. The higher the socioeconomic disparity, the lower the municipal bonds are ultimately rated. As Chen et. al. states:

“The better a jurisdiction’s economic status, the higher the potential revenues the jurisdiction can generate, and the lower the probability that the bonds issued by the jurisdiction would default.”^17^

This paints a clear picture of the cycle local governments with lower socioeconomic measures find themselves in. In today’s macroeconomics environment, interest rates are on the rise. Rising interest rates have also been shown to disproportionately affect lower rated borrowers^20^, which is further detrimental to county’s with low credit ratings and their populations. Poor infrastructures have been shown to be associated with reduced credit ratings on a state level^17^, which may additionally contribute to the cycle governments with low credit ratings may find themselves in.

It should be noted that this study is, by definition, an observational study and therefore cannot establish any causal relationship between county credit ratings and mortality rates. More research, specifically experimental research, is needed in order to definitively state that increasing county credit ratings could lead to improvements in public health outcomes.

Overall, the findings of this study highlight the importance of various financial factors in the understanding and correction of public health outcome disparities. The implications of this research may be considered by both policymakers and public health researchers alike in determining the optimal way to support local municipalities financially. Further research can be undertaken from here to help understand the interlinking of local bond markets and their ultimate effect on public health, and policy may be designed to mitigate or assist with the larger costs of borrowing low socioeconomic status communities face. Understanding these factors and their effect on inequality and cycles of poverty is critical in order to ultimately improve the health of a country’s citizens. It is our hope that using an interdisciplinary research approach to guide policy will help alleviate disparities in both Michigan communities, as well as communities around the globe.

## Limitations

The association found between county credit ratings and age-adjusted county mortality rates, although significant, does not necessarily imply causation. There are many possible confounding variables detrimental to the study hypothesis. For instance, some states offer protections to local governments in danger of defaulting, which may skew county credit ratings higher and borrowing rates lower^21^. Additionally, debt and taxation policies surrounding the local and broader state governments can change county credit ratings significantly. For instance, restrictions on state debt issuance have been shown to lead to higher credit ratings, and revenue limits (i.e. taxation limits) have been shown to lead to higher interest rates through lower credit ratings^22^.

Only counties located in Michigan were researched and not all counties in Michigan have active credit ratings on Moodys. Specifically, rural counties with smaller populations were the bulk of the counties without credit ratings. This is likely due to rural counties both issuing reduced quantities of public bonds, and/or not always paying for a bond rating company to assess the bond. This not only reduced our sample size within Michigan, but left out the majority of county governments, which, if able to be included, may have influenced the results.

Another potential confounding variable is that counties may contain areas with large spreads of wealth. For example, Wayne county contains both the city of Grosse Pointe Farms, which has an average income of $146,667, and the city of Detroit, which has an average income of $34,762^23^. These large distributions in wealth may have the effect of skewing the resultant rating for a specific county.

Lastly, political instability in a local government may negatively impact its credit rating. This can cause disruptions in data, as a county may have a short-lived bout of political turmoil which subsequently hurts their credit rating, despite the mortality measures remaining the same^**24**^.

## Supporting information

Data Analysis Supplemental Information

## Data Availability

All data used in the study was ascertained from the following URLs:
https://www.moodys.com/researchandratings/region/north-america/michigan/03F028?tb=0&ol=-1&lang=en
https://www.mdch.state.mi.us/osr/CHI/Prov/ProvisionalQuarterlyReported.asp
https://www.census.gov/quickfacts/fact/table/detroitcitymichigan,grossepointefarmscitymichigan,grossepointecitymichigan,MI/INC110221

https://www.moodys.com/researchandratings/region/north-america/michigan/03F028?tb=0&ol=-1&lang=en

https://www.mdch.state.mi.us/osr/CHI/Prov/ProvisionalQuarterlyReported.asp

https://www.census.gov/quickfacts/fact/table/detroitcitymichigan,grossepointefarmscitymichigan,grossepointecitymichigan,MI/INC11022

## Footnotes

### Contributors

Both authors provided the design of the study. conducted the data analysis. Both authors provided large contributions to the interpretation of the results and drafting of the paper. Both authors are guarantors.

### Sponsors

None declared.

### Ethical approval

Not required.

### Data Availability Statement

All data used in the study was ascertained from the following URLs:

- https://www.moodys.com/researchandratings/region/north-america/michigan/03F028?tb=0&ol=-1&lang=en
- https://www.mdch.state.mi.us/osr/CHI/Prov/ProvisionalQuarterlyReported.asp
- https://www.census.gov/quickfacts/fact/table/detroitcitymichigan,grossepointefarmscitymichigan,grossepointecitymichigan,MI/INC110221

### Transparency declaration

The authors affirms that this manuscript is an honest, accurate, and transparent account of the study being reported; that no important aspects of the study have been omitted; and that any discrepancies from the study as planned (and, if relevant, registered) have been explained

