## Supplementary material for "An Ecological Study on the Associations of County-Based Credit Ratings with Age-Adjusted Mortality Rates in Michigan": Data Analysis Supplemental Information

| *All Mortality data gathered from https://www.mdch.state.mi.us/osr/CHI/Prov/ProvisionalQuarterlyReported.asp | County | Rating (Moodys) | Grouping (For Regression ) | Mortality (Q1 2022) - Age Adjusted | Mortality (Q4 2021) - Age Adjusted | Mortality (Q3 2021) - Age Adjusted | Mortality (Q2 2021) - Age Adjusted | Mortality (Q1 2021) - Age Adjusted | Mortality (Q4 2020) - Age Adjusted |
|---|---|---|---|---|---|---|---|---|---|
| | Kent | Aaa | 1 | 824.2 | 812.7 | 810.1 | 806.3 | 809.5 | 798.3 |
| | Livingston | Aaa | 1 | 827.1 | 811.2 | 783.8 | 757.9 | 762.9 | 748.7 |
| | Oakland | Aaa | 1 | 776.9 | 765.1 | 754.7 | 752.6 | 792.8 | 794.5 |
| | Ottawa | Aaa | 1 | 779.8 | 753.5 | 756.5 | 737.3 | 748.7 | 734.7 |
| | Washtenaaw | Aaa | 1 | 703.2 | 692 | 692 | 689 | 703.6 | 701.8 |
| | Clinton | Aa1 | 2 | 802.4 | 802.1 | 798.3 | 808.3 | 800.4 | 777.8 |
| | Macomb | Aa1 | 2 | 958.7 | 938.7 | 911.5 | 901.4 | 958 | 952.5 |
| | Saginawa | Aa2 | 3 | 980.4 | 972.5 | 1012.6 | 1015.8 | 1035 | 1006.4 |
| | Emmett | Aa2 | 3 | 747.8 | 739 | 1040.9 | 1045.5 | 997.2 | 979.2 |
| | St Claire | Aa2 | 3 | 1068.1 | 1070.2 | 954.7 | 958.5 | 925.6 | 920.5 |
| | Lenawee | Aa2 | 3 | 1021.4 | 979.2 | 1073.8 | 1071.5 | 1059.5 | 993.8 |
| | Bay | Aa2 | 3 | 1017.6 | 1027.2 | 803.3 | 805 | 788.9 | 772.8 |
| | Grand Travers | Aa2 | 3 | 845.7 | 823.8 | 804.3 | 836.9 | 850.1 | 829.7 |
| | Inhgam | Aa2 | 3 | 971.1 | 943.5 | 935.1 | 908.2 | 880.5 | 874.1 |
| | Berrien | Aa2 | 3 | 947.3 | 902.1 | 884.3 | 867.8 | 889.1 | 856.7 |
| | Barry | Aa2 | 3 | 972.6 | 963.3 | 896 | 857.8 | 854.5 | 801.4 |
| | Jackson | Aa3 | 4 | 1035.1 | 1024.9 | 1006.5 | 991.8 | 982.1 | 958 |
| | Alpena | Aa3 | 4 | 958.5 | 968.2 | 867.2 | 884.7 | 832.9 | 835.8 |
| | Crawford | A1 | 5 | 977.5 | 977.2 | 913.5 | 865.2 | 841 | 756.1 |
| | Genesee | A1 | 5 | 1120.6 | 1077.2 | 1063.4 | 1046.9 | 1066.6 | 1044.6 |
| | Wayne | A1 | 5 | 1069.5 | 1048.5 | 1006.5 | 1008.8 | 1102 | 1107.4 |

### REGRESSION ANALYSIS - OUTPUT ON SHEET 2

**SUMMED 2020 Q4 - 2022 Q1**

| | |
|---|---|
| 1 | 810.18333 |
| 1 | 781.93333 |
| 1 | 772.76667 |
| 1 | 751.75 |
| 1 | 696.93333 |
| 2 | 798.21667 |
| 2 | 936.8 |
| 3 | 1003.7833 |
| 3 | 924.93333 |
| 3 | 982.93333 |
| 3 | 1033.2 |
| 3 | 869.13333 |
| 3 | 831.75 |
| 3 | 918.75 |
| 3 | 891.21667 |
| 3 | 890.93333 |
| 4 | 999.73333 |
| 4 | 891.21667 |
| 5 | 888.41667 |
| 5 | 1069.8833 |
| 5 | 1057.1167 |

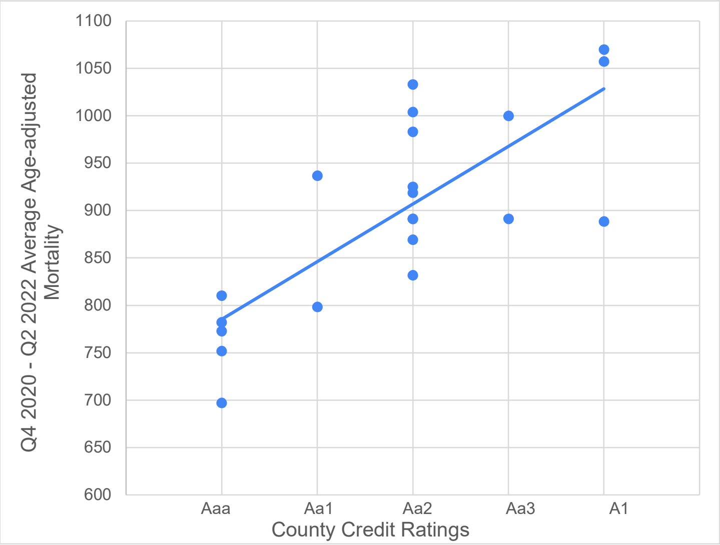


**Figure 1**

SUMMARY OUTPUT

| *Regression Statistics* | |
|---|---|
| Multiple R | 0.771544866 |
| R Square | 0.595281481 |
| Adjusted R Square | 0.573980506 |
| Standard Error | 68.2222669 |
| Observations | 21 |

ANOVA

| | *df* | *SS* | *MS* | *F* | *Significance F* |
|---|---|---|---|---|---|
| Regression | 1 | 130069.42 | 130069.415 | 27.94620852 | 4.21007E-05 |
| Residual | 19 | 88431.276 | 4654.2777 | | |
| Total | 20 | 218500.69 | | | |

| | *Coefficients* | *Standard Error* | *t Stat* | *P-value* | *Lower 95%* | *Upper 95%* | *Lower 95.0%* | *Upper 95.0%* |
|---|---|---|---|---|---|---|---|---|
| Intercept | 724.6211261 | 35.555625 | 20.3799295 | 2.26144E-14 | 650.2023481 | 799.0399042 | 650.2023481 | 799.0399042 |
| X Variable 1 | 60.75490991 | 11.492643 | 5.28641736 | 4.21007E-05 | 36.7005307 | 84.80928912 | 36.7005307 | 84.80928912 |
